# Evaluation of patients treated by telemedicine in the COVID-19 pandemic by a private clinic in São Paulo, Brazil: A non-randomized clinical trial preliminary study

**DOI:** 10.1101/2021.11.05.21265569

**Authors:** Michelle Chechter, Gustavo Maximiliano Dutra da Silva, Thomas Gabriel Miklos, Marta Maria Kemp, Nilzio Antonio da Silva, Gabriel Lober, Marcela Ferreira Tavares Zanut, Rute Alves Pereira e Costa, Aline Pinheiro dos Santos Cortada, Luciana de Nazare Lima da Cruz, Paulo Macio Porto de Melo, Bruno Campello de Souza, Morton Aaron Scheinberg

## Abstract

**Introduction:** As a result of the coronavirus disease 2019 (COVID-19) pandemic the year 2020 brought major changes on the delivery of health care and face to face physician patient communication was significantly reduced and the practice of remote telehealth care using computer technology is assuming a standard of care, particularly, with COVID-19 patients with attempts to reduce viral spread.

**Objective:** To describe the clinical practice experience using telemedicine towards COVID-19 and the respective clinical outcomes.

**Methods:** We performed a pilot open-label non-randomized, controlled clinical trial. The patients were divided into four groups, according severity of symptoms: (1) asymptomatic, (2) mild symptoms, (3) moderate symptoms and (4) severe symptoms, and were followed up for five days, counted from the beginning of the symptoms. A drug intervention was performed in group 3, for which the protocol followed as suggested by the International Pulmonology Society’s consensus for adults with moderate symptoms: first day (attack phase) hydroxychloroquine sulfate 400 mg 12/12h; second to fifth day (maintenance phase) 200 mg (half pill) 12/12h. The medication was associated with azithromycin 500mg once a day for five days. For children with moderate symptoms were used: hydroxychloroquine sulfate 6.5 mg/kg/dose every 12 hours in the first day and 3.25 mg/kg/dose every 12 hours from day 2 to 5. The therapeutic response was telemonitored. Group 4 patients were directly oriented to seek hospital care. During the use of the drugs, the patients were telemonitored daily.

**Results:** One hundred eighty-seven patients were seen with mean age of 37,6 years (±15,6). The most frequent symptom was cough (57,6%), followed by malaise (60,3%), fever (41,1%), headache (56,0%), muscle pain (51,1%). Of all the patients that sought telemedicine service in our center, 23% were asymptomatic despite contact with people with probable diagnostic of COVID-19; 29,4% reported mild symptoms, 43,9% moderate symptoms, and 3,7% severe symptoms. It was possible to observe in patients treated their symptoms of COVID-19 (group 3) with hydroxychloroquine and azithromycin for five days, presented statistically better improvement of the symptoms when compared to those that did not follow the protocol (p = 0.039). Three patients were hospitalized and discharged after recovery.

**Conclusions:** Our study showed that patients with COVID-19 who had delivery of health care through telemedicine initiated in early stages of the disease presented satisfactory clinical response, reducing the need of face-to-face consultations and hospitalizations. Our results indicate that the use of telemedicine with diagnosis and drug treatment protocols is a safe and effective strategy to reduce overload of health services and the exposure of healthcare providers and the general population to infected patients in a pandemic situation.

**Trial registration:** RBR-658khm

Human Research Ethics Committee number: 30246520.0.0000.0069

## Introduction

The pandemic the year 2020 brought major changes on the delivery of health care and face to face physician patient communication was significantly reduced and the practice of remote telehealth care through the use of computer technology is assuming a standard of care, particularly, with COVID-19 patients with attempts to reduce viral spread [1,2,3,4]. In the present study we describe the clinical experience using telemedicine towards COVID-19 and evaluate the respective clinical outcomes. It was performed during the first semester of 2020 giving advice and treatment with confirm diagnosis or suspect or having the disease in an attempt to reduce face to face service, exposure to health care workers and eventual need of hospitalization.

## Methods

This is an open-label non-randomized, controlled clinical trial, initiated after pandemic state and quarantine had been declared by the government of the state of São Paulo, where Mazzei’s Medical Center (MMC) started a telemedicine service, as regulated by the Health Ministry (HM) and Federal Council of Medicine (FCM), for the care of patients who presented with symptoms that could be related to COVID-19.The consultations were performed, as suggested by the American Academy of Allergy Asthma and Immunology [2], by physicians that are registered in the Regional Council of Medicine of the State of São Paulo and present a Specialist Qualification Record. All interviews occurred though video calls using the platform *WhatsApp Facebook Inc*, as recommended for the practice of Telemedicine by the *American Academy of Allergy Asthma and Immunology* (Figure/ flow diagram I). In the interview, we used a semi-structured questionnaire developed by the authors (supplemental I). The data was recorded in a certified medical record DFMed, remotely accessed through *AnyDesk* platform and, posteriorly, tabulated and stored with *Microsoft Excel 2003* software. We have considered as inclusion criteria patients who sought our service for medical care for diagnosis, treatment or clarifying doubts related do COVID-19 There were no restrictions for age or sex.

For home treatment, according to the developed protocol, the eligible patients had clinical diagnosis of COVID-19 with suggestive symptoms, progressive worsening of the symptoms and who had had previous contact with suspected or laboratory (RT-PCR) confirmed case source. Suggestive symptoms considered were persistent coughing, fever, muscle pain, headache, diarrhea, chills, anorexia, important weight loss, conjunctival irritation, anosmia and dyspnea of abrupt onset with progressive worsening [5].

The patients were divided into four groups: (1) asymptomatic, (2) mild symptoms (those who referred clinical improvement with the use of symptomatic or antibiotic therapy not specific for the treatment of infection by SARS-CoV-2, and who didn’t need other medical interventions), (3) moderate symptoms (patients with more intense and progressive symptoms such as fever (temperature greater than or equal to 37,8 Celsius degrees), persistent coughing, anosmia, mild muscle pain, mild dyspnea, persistent diarrhea, which didn’t improve with symptomatic medication) and (4) severe symptoms (patients with even more intense and progressive symptoms as those in group 3) (supplemental II).

Symptomatic patients were followed for 5 days counted from the first telemedicine interview. Medical intervention directed to treatment of COVID-19 was proposed only in group 3, for which the drug protocol was prescribed according to the International Pulmonology Society’s consensus [5] for adults with moderate symptoms: first day (attack phase) hydroxychloroquine sulfate 400 mg 12/12h; second to fifth day (maintenance phase) 200 mg (half pill) 12/12h. The medication was associated with azithromycin 500 mg once a day for five days. For children with moderate symptoms were used: hydroxychloroquine sulfate 6.5 mg/kg/dose in the first day and 3.25 mg/kg/dose every 12 hours from day 2 to 5. The therapeutic response was telemonitored. Group 4 patients were directly oriented to seek hospital care. During the use of the drugs, the patients were telemonitored daily. Patients with moderate symptoms (group 3) that did not accepted the treatment with hydroxychloroquine were included in the study as controlled group.

Treatment was evaluated in all symptomatic patients by a clinical improvement scale on the 5^th^ day of therapy. At this day patients were interviewed by a physician blinded to the other procedures of study just to apply the clinical improvement score.

We have evaluated cardiologic contraindication for the use of hydroxychloroquine through the risk stratification of the American College of Cardiology [6] (supplemental III).

All patients received the medications hydroxychloroquine sulfate compound and azithromycin marketed in Brazil by the regulatory national agency. Also patients received systematic orientations for disinfection of the family through ambient, objects and contact surfaces hygiene, cleaning (sprinkling) with hydrated ethanol 70° INPM and door sealing in condominiums. Personal hygiene when coming in and going out of the family nucleus and the use of equipment for individual protection (such as mask) was reinforced. After receiving all information about the study, patients accepting to participate in the project signed the informed consent, and all regulatory requirements for the execution of such a research in Brazil were met.

## Statistical analysis

Qualitative characteristics of the patients were described using absolute and relative frequency and quantitative characteristics were described using summary measures (mean, standard deviation, minimum and maximum) for all the sample. Clinical improvement percentage and adverse effects occurrence were presented for all patients with moderate symptoms that followed the protocol treatment.

The qualitative characteristics were described according to the severity of symptoms and the association was verified using likelihood ratio test [7], ages were compared using analysis of variances (ANOVA) and the number of people living in the residency of the patients and the number of those presenting symptoms were compared according to the severity of symptoms using Kruskal-Wallis tests followed by Dum’s multiple comparisons to verify among which severity of symptoms there were differences.

Of the patients with mild and moderate symptoms, the qualitative characteristics were described according to the COVID-19 specific treatment and the association was verified using chi-square test or exact tests (Fisher’s exact test or likelihood ratio test) [7] and the ages were compared according to the treatment using t-Student test [7], and the number of people living in the residency of the patients and the number of those presenting symptoms were compared using Mann-Whitney tests [7].

For the statistical analysis, the statistical package *StatistiXL* was used (Statistical Power for MS Excel version 1.8, 2007). The analysis was performed with the IBM-SPSS software for Windows version 20.0 (IBM, IL, Chicago, EUA). The tests were made with a level of significance of 5%.

## Results

On table 1 we present the demographics of the study. The average age was 37.6 (±15.6). More than half (64.3%) lived with a spouse or companion and 78.1% declared to be white, 15.3% brown and 6.6% black. In relation to personal history, 29.9% referred to have had a previous surgery and 48.6% to have had some chronic previous or current illness of which 15.1% were respiratory, 51.4% used some medication daily, 5.6% referred to be smokers, 20.3% would take alcohol in a social context, 9.6% declared to be alcoholics and 1.1% declared to use some kind of illicit drug. The average number of people cohabiting the symptomatic patient’s houses was 3,35 and the average of other family members with symptoms was 1.27±0.99 (table 1).

**Table 1.** Description of population’s characteristics and patients clinical history, mean, standard deviation, median, minimum and maximum of patients evaluated. MMC, 2020.

| Variable | Description<br>(N- 187) |
| --- | --- |
| Age (years) |  |
| average ± SD | 37,6 ± 15,6 |
| median (min.; max.) | 36,8 (0,9; 84,8) |
| Race/color |  |
| White | 143 (78,1%) |
| Brown | 28 (15,3%) |
| Black | 12 (6,6%) |
| Marital Status |  |
| Single | 55 (29,7%) |
| Married | 119 (64,3%) |
| Divorced | 8 (4,3%) |
| Widow | 3 (1,6%) |
| Religion |  |
| Catholic | 93 (52,8%) |
| Evangelical | 27 (15,3%) |
| Jewish | 7 (4%) |
| Spirits | 22 (12,5%) |
| Atheist | 4 (2,2%) |
| Agnost | 15 (8,5%) |
| Other | 8 (4,5%) |
| Previous surgeries | 52/174 (29,9%) |
| Clinical previous or current illness | 86/187 (48,6%) |
| Respiratory disease | 25/166 (15,1%) |
| N. of people living in the same residency |  |
| average ± SD | 3,35 ± 1,29 |
| median (min.; max.) | 3 (1; 9) |
| N. of people in the same residency with symptoms |  |
| average ± SD | 1,27 ± 0,99 |
| median (min.; max.) | 1 (0; 4) |
| Medications in use | 89/173 (51,4%) |
| Smoker | 10 (5,6%) |
| Alcoholism |  |
| No | 124 (70,1%) |
| Social | 36 (20,3%) |
| Yes | 17 (9,6%) |
| Drug addiction | 2/174 (1,1%) |

SD: standard deviation; min: minimum; max: maximum; N.: number

In this study, 187 patients were analyzed: 43 asymptomatic and 144 symptomatic classified according to the severity of their symptoms as showed in figure 1. Of these patients, 55 presented with mild symptoms of which only one evolved with abrupt deterioration and was referred for hospital treatment; 74 presented with moderate symptoms and were offered the protocol treatment for COVID-19 with hydroxychloroquine and azithromycin, 60 adhered to treatment (intervention group) and 12 did not accepted hydroxychloroquine, choosing be treated only with azithromycin (control group); 3 presented with severe symptoms and were immediately referred to the hospital. One patient in the severe group diagnosed with viral myocarditis developed tachycardia during hospital treatment, the medications (hydroxychloroquine and azithromycin) were stopped and the tachycardia remained until the total improvement of the symptoms.

**Figure 1.**
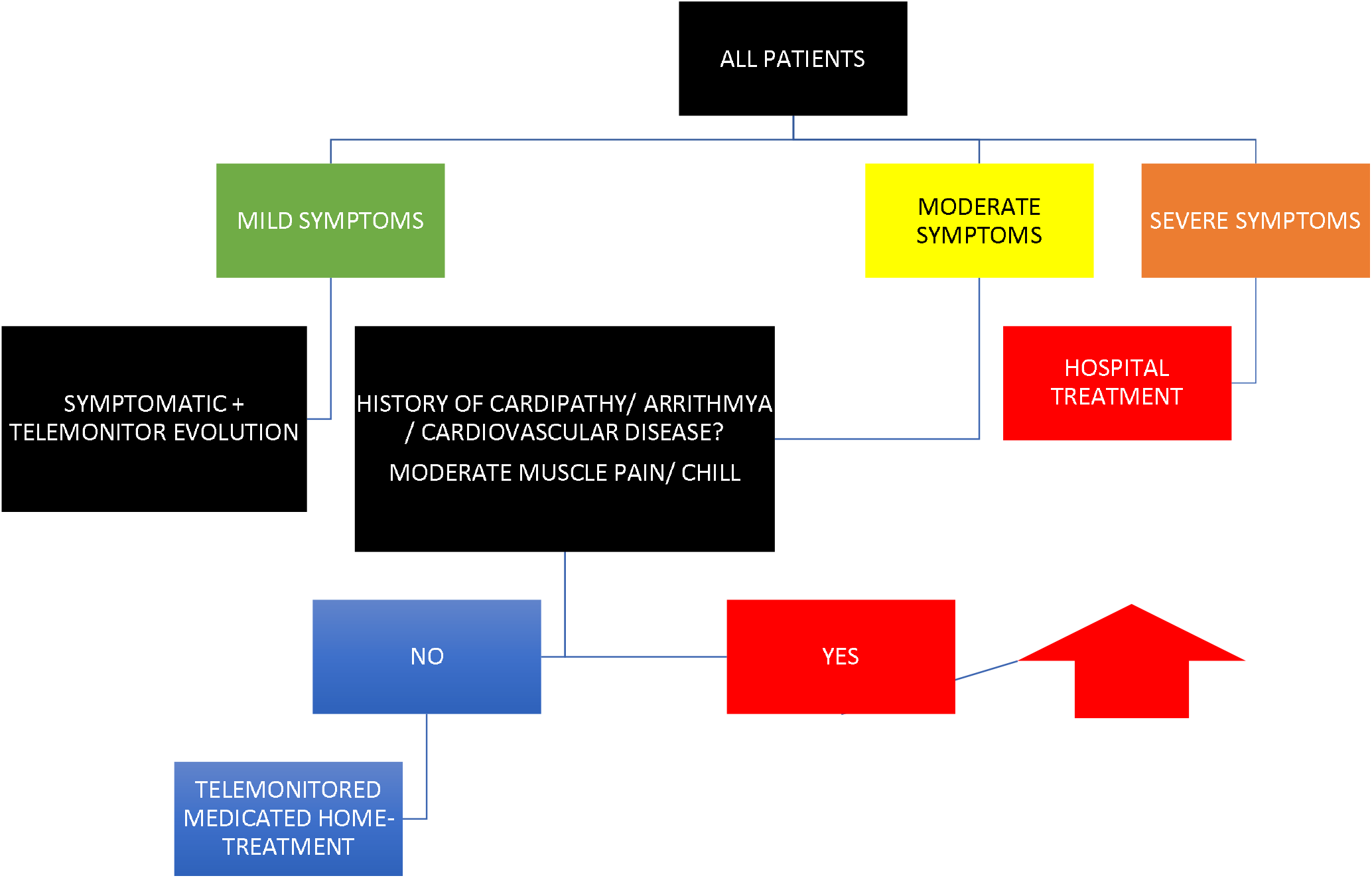
Screening flow diagram for Telemedicine – Mazzei’s Medical Center, 2020.

Nine out twelve patients with moderate symptoms of the control group presented progressive worsening of the symptoms and were referred to the hospital, resulting in 3 hospitalizations. Of those who followed the protocol (intervention group), all 60 had improvement of their symptoms and, during telemonitored treatment, had no need to be referred to hospitals. It is noticeable that one patient from the intervention group only had improvement of the symptoms after suspension of the medication of continuous use Dexlanzoprazol, a proton-pump inhibitor. Inadequate treatment was considered in 12 patients (control group) who were afraid of using hydroxychloroquine and chose to use only azithromycin.

The average age was 37.6 (±15.6). More than half (64.3%) lived with a spouse or companion and 78.1% declared to be white, 15.3% brown and 6.6% black. In relation to personal history, 29.9% referred to have had a previous surgery and 48.6% to have had some chronic previous or current illness of which 15.1% were respiratory, 51.4% used some medication daily, 5.6% referred to be smokers, 20.3% would take alcohol in a social context, 9.6% declared to be alcoholics and 1.1% declared to use some kind of illicit drug. The average number of people cohabiting the symptomatic patient’s houses was 3,35 and the average of other family members with symptoms was 1.27±0.99 (table 1).

On table 2 we describe the most common symptoms during presentation, indisposition, indisposition (60,5%) followed by cough (57.6%), muscle pain (51,1%), anosmia (45,9%), decreased appetite (44,5%), sore throat (38,6%), diarrhea (28%), conjunctival irritation (20%), weight loss (19,2%), vomit (10,5%), and blood in the stool (2,4%). Of all patients, 46,4% referred improvement of symptoms with more frequent baths during the day, 64,9% with the intake of dipyrone or acetaminophen and 60% were feeling well at the moment of the interview.

**Table 2.** General description of clinical symptoms of patients who sought our telemedicine service. MMC, 2020.

| **Variable** | **Description**<br>**N=144** |
| --- | --- |
| Indisposition | 111/184 (60,5%) |
| Cough | 106/184 (57,6%) |
| Muscle pain | 94/184 (51,1%) |
| Improvement of symptoms with bath* | 54/116 (46,4%) |
| Anosmia | 62/135 (45,9%) |
| Decreased appetite | 81/182 (44,5%) |
| Sore throat | 71/184 (38,6%) |
| Diarrhea | 51/182 (28%) |
| conjunctival irritation | 34/170 (20%) |
| Weight loss | 30/156 (19,2%) |
| Vomit | 19/181 (10,5%) |
| Blood in the stool | 4/168 (2,4%) |

\* Only for applicable cases

On table 3 we present found that 52,7% of the patients had face-to-face contact with people presenting flu-like symptoms, 41,8% with people with RT-PCR positive test for COVID-19 and 9% had recently travelled. The 3 patients in group 4 (severe symptoms) took the RT-PCR for COVID-19 test and presented positive results.

**Table 3.** Description of epidemiologic criteria evaluated and/or laboratory confirmation for diagnostic screening of COVID-19 for patients who sought our telemedicine service. MMC, 2020.

| **Variable** | **Description**<br>**N=187** |
| --- | --- |
| Contact with suspected case | 97 (52,7%) |
| Contact with confirmed source case | 77 (41,8%) |
| Recent travel | 16/177 (9%) |
| Positive laboratory test for COVID-19 | 40/187 (21,4%) |

In table 4 the status of the clinical presentation is presented, 23% of the patients who sought our telemedicine service were asymptomatic, 29,4% had mild symptoms, 43,9% had moderate symptoms and 3.7% severe symptoms.

**Table 4.** Staggering of group of patients divided according to severity of symptoms of COVID-19, for the application of our home-treatment protocol. MMC, 2020.

| **Variable** | **Description**<br>**N=187** |
| --- | --- |
| Group |  |
| Asymptomatic | 43 (23%) |
| Mild symptoms | 55 (29,4%) |
| Moderate symptoms | 82 (43,9%) |
| Severe symptoms | 7 (3,7%) |

It was possible to observe a statistically significant association with the groups of severity of cough symptoms, weight loss, muscle pain and decreased smell (p <0.05), with the frequency of these symptoms being higher in the most severe patients.

Table 5 shows the distribution of related flu-like symptoms according to the severity of clinical symptoms and statistical tests results.

**Table 5.** Description of distriubution of related to flu-like symptoms according to the severity of clinical symptoms and statistical tests results. MMC, 2020.

| Variable | Group |  |  | p |
| --- | --- | --- | --- | --- |
|  | Mild symptoms | Moderate symptoms | Severe symptoms |  |
| **Age (years)** |  |  |  | 0,356* |
| average ± SD | 38,7 ± 16,7 | 37,5 ± 15,3 | 46,3 ± 9,4 | |
| median (min.; max.) | 37 (0,9; 84,8) | 38,6 (1,4; 73) | 44,8 (34; 61,9) |  |
| **Previous surgeries, n (%)** |  |  |  | 0,184 |
| No | 33 (70,2%) | 56 (70,9%) | 2 (33,3%) |  |
| Yes | 14 (29,8%) | 23 (29,1%) | 4 (66,7%) |  |
| **Other clinical previous or current illness, n (%)** |  |  |  | 0,426 |
| No | 36 (80%) | 62 (79,5%) | 4 (57,1%) |  |
| Yes | 9 (20%) | 12 16 (20,5%) | 3 (42,9%) |  |
| **Respiratory disease, n (%)** |  |  |  | 0,982 |
| No | 36 (83,7%) | 64 (83,1%) | 6 (85,7%) |  |
| Yes | 7 (16,3%) | 13 (16,9%) | 1 (14,3%) |  |
| **Medications in use, n (%)** |  |  |  | 0,302 |
| No | 23 (51,1%) | 1 (38,8%) | 2 (28,6%) |  |
| Yes | 22 (48,9%) | 49 (61,3%) | 5 (71,4%) |  |
| **Smoker, n (%)** |  |  |  | 0,297 |
| No | 45 (95,7%) | 77 (95%) | 7 (100%) |  |
| Yes | 2 (4,3%) | 4 (4,9%) | 0 (0) |  |
| **Alcoholism, n (%)** |  |  |  | 0,717 |
| No | 31 (66%) | 55 (67,9%) | 6 (85,7%) |  |
| Social | 11 (23,4%) | 17 (21%) | 1 (14,3%) |  |
| Yes | 5 (10,6%) | 9 (11,1%) | 0 (0%) |  |
| **N. of people living in the same residency** |  |  |  | 0,644£ |
| average ± SD | 3,46 ± 1,66 | 3,41 ± 1,08 | 3,14 ± 1,22 | |
| median (min.; max.) | 3 (1; 9) | 3 (1; 9) | 4 (1; 4) |  |
| **N. of people in the same residency with symptoms** |  |  |  | 0,306£ |
| average ± SD | 1,48 ± 0,8 | 1,69 ± 0,9 | 1,57 ± 0,79 | |
| median (min.; max.) | 1 (0; 4) | 2 (0; 4) | 1 (1; 3) |  |
| **Fever, n (%)** |  |  |  | 0,530 |
| No | 28 (51,9%) | 36 (43,9%) | 2 (33,3%) |  |
| Yes | 26 (48,1%) | 46 (56,1%) | 4 (66,7%) |  |
| **Cough, n (%)** |  |  |  | 0,033 |
| No | 18 (34,6%) | 18 (22%) | 0 (0%) |  |
| Yes | 34 (65,4%) | 64 (78%) | 7 (100%) |  |
| **Nasal congestion, n (%)** |  |  |  | 0,611 |
| No | 25 (49%) | 33 (40,2%) | 3 (42,9%) |  |
| Yes | 26 (51%) | 49 (59,8%) | 4 (57,1%) |  |
| **Diarrhea, n (%)** |  |  |  | 0,482 |
| No | 34 (66,7%) | 51 (63%) | 3 (42,9%) |  |
| Yes | 17 (33,3%) | 30 (37%) | 4 (57,1%) |  |
| **Blood in the stool, n (%)** |  |  |  | 0,138 |
| No | 44 (100%) | 73 (94,8%) | 4 (100%) |  |
| Yes | 0 (0%) | 4 (5,2%) | 0 (0%) |  |
| **Conjunctival irritation, n (%)** |  |  |  | 0,062 |
| No | 38 (84,4%) | 52 (68,4%) | 3 (50%) |  |
| Yes | 7 (15,6%) | 24 (31,6%) | 3 (50%) |  |
| **Vomit, n (%)** |  |  |  | 0,103 |
| No | 47 (94%) | 66 (81,5%) | 6 (85,7%) |  |
| Yes | 3 (6%) | 15 (18,5%) | 1 (14,3%) |  |
| **decreased appetite, n (%)** |  |  |  | **0,006** |
| No | 30 (58,8%) | 25 (30,9%) | 3 (42,9%) |  |
| Yes | 21 (41,2%) | 56 (69,1%) | 4 (57,1%) |  |
| **Weight loss, n (%)** |  |  |  | 0,007 |
| No | 34 (89,5%) | 47 (67,1%) | 2 (40%) |  |
| Yes | 4 (10,5%) | 23 (32,9%) | 3 (60%) |  |
| **Shortness of breath, n (%)** |  |  |  | 0,496 |
| No | 33 (64,7%) | 44 (54,3%) | 4 (57,1%) |  |
| Yes | 18 (35,3%) | 37 (45,7%) | 3 (42,9%) |  |
| **Indisposition, n (%)** |  |  |  | 0,109 |
| No | 14 (26,9%) | 16 (19,5%) | 0 (0%) |  |
| Yes | 38 (73,1%) | 66 (80,5%) | 7 (100%) |  |
| **Muscle pain, n (%)** |  |  |  | 0,014 |
| No | 23 (43,4%) | 24 (29,6%) | 0 (0%) |  |
| Yes | 30 (56,6%) | 57 (70,4%) | 7 (100%) |  |
| **Headache, n (%)** |  |  |  | 0,055 |
| No | 20 (39,2%) | 17 (21%) | 1 (14,3%) |  |
| Yes | 31 (60,8%) | 64 (79%) | 6 (85,7%) |  |
| **Sore throat, n (%)** |  |  |  | 0,205 |
| No | 22 (41,5%) | 46 (56,8%) | 3 (42,9%) |  |
| Yes | 31 (58,5%) | 35 (43,2%) | 4 (57,1%) |  |
| **Anosmia, n (%)** |  |  |  | 0,001 |
| No | 33 (64,7%) | 40 (51,9%) | 0 (0%) |  |
| Yes | 18 (35,3%) | 37 (48,1%) | 7 (100%) |  |
| **Improvement of symptoms with bath\*** |  |  |  | 0,457 |
| No | 22 (61,1%) | 37 (50%) | 2 (40%) |  |
| Yes | 14 (38,9%) | 37 (50%) | 3 (60%) |  |

Likelihood ratio test; \* ANOVA; £ Kruskal-Wallis test

On table 6 we show the description of populational and clinical characteristics related to clinical symptoms of the patients presenting moderate symptoms and received the protocol treatment for COVID-19 with hydroxychloroquine and azithromycin, related to the patients who took only azithromycin, and the respective statistical tests results.

**Table 6.** Patients with moderate symptoms and treatment administered for respective groups hydroxychloroquine and azithromycin and only azithromycin.

| Variable | Protocol treatment for COVID-19 |  | p |
| --- | --- | --- | --- |
|  | No (N = 7) | Yes (N = 75) |  |
| **Age (years)** |  |  | 0,083<sup>b</sup> |
| average ± SD | 47,1 ± 8,5 | 36,6 ± 15,5 | |
| median (min.; max.) | 47,8 (32,1; 57,6) | 37,5 (1,4; 73) |  |
| **Previous surgeries, n (%)** |  |  | 0,409<sup>a</sup> |
| No | 4 (57,1%) | 52 (72,2%) |  |
| Yes | 3 (42,9%) | 20 (27,8%) |  |
| **Clinical previous or current illness, n (%)** |  |  | 0,628 |
| No | 5 (71,4%) | 57 (80,3%) |  |
| Yes | 2 (28,6%) | 14 (19,7%) |  |
| **Respiratory or imunossupressor illnesses, n (%)** |  |  | >0,999 |
| No | 6 (85,7%) | 58 (82,9%) |  |
| Yes | 1 (14,3%) | 12 (17,1%) |  |
| **Medication in use, n (%)** |  |  | 0,239 |
| No | 1 (14,3%) | 30 (41,1%) |  |
| Yes | 6 (85,7%) | 43 (58,9%) |  |
| **Smoker, n (%)** |  |  | 0,627# |
| No | 7 (100%) | 69 (94,6%) |  |
| Yes | 0 (0%) | 4 (5,4%) |  |
| **Alcoholism, n (%)** |  |  | 0,057# |
| No | 7 (100%) | 48 (64,9%) |  |
| Social | 0 (0%) | 17 (23%) |  |
| Yes | 0 (0%) | 9 (12,2%) |  |
| **N. of people living in the same residency** |  |  | 0,502£ |
| average ± SD | 3,43 ± 1,4 | 3,41 ± 1,05 | |
| median (min.; max.) | 4 (1; 5) | 3 (1; 9) |  |
| **N. of people in the same residency with symptoms** |  |  | 0,305£ |
| average ± SD | 1,29 ± 0,76 | 1,73 ± 0,91 | |
| median (min.; max.) | 1 (0; 2) | 2 (0; 4) |  |
| **Fever, n (%)** |  |  | 0,231 |
| No | 5 (71,4%) | 31 (41,3%) |  |
| Yes | 2 (28,6%) | 44 (58,7%) |  |
| **Cough, n (%)** |  |  | >0,999 |
| No | 1 (14,3%) | 17 (22,7%) |  |
| Yes | 6 (85,7%) | 58 (77,3%) |  |
| **Nasal congestion, n (%)** |  |  | >0,999 |
| No | 3 (42,9%) | 30 (40%) |  |
| Yes | 4 (57,1%) | 45 (60%) |  |
| **Diarrhea, n (%)** |  |  | >0,999 |
| No | 5 (71,4%) | 46 (62,2%) |  |
| Yes | 2 (28,6%) | 28 (37,8%) |  |
| **Blood in the stool, n (%)** |  |  | 0,322 |
| No | 6 (85,7%) | 67 (95,7%) |  |
| Yes | 1 (14,3%) | 3 (4,3%) |  |
| **Conjunctival irritation, n (%)** |  |  | >0,999 |
| No | 4 (66,7%) | 48 (68,6%) |  |
| Yes | 2 (33,3%) | 22 (31,4%) |  |
| **Vomit, n (%)** |  |  | >0,999 |
| No | 6 (85,7%) | 60 (81,1%) |  |
| Yes | 1 (14,3%) | 14 (18,9%) |  |
| **Reduction of appetite, n (%)** |  |  | 0,670 |
| No | 3 (42,9%) | 22 (29,7%) |  |
| Yes | 4 (57,1%) | 52 (70,3%) |  |
| **Weight loss, n (%)** |  |  | 0,656 |
| No | 5 (83,3%) | 42 (65,6%) |  |
| Yes | 1 (16,7%) | 22 (34,4%) |  |
| **Shortness of breath, n (%)** |  |  | 0,404 |
| No | 2 (33,3%) | 42 (56%) |  |
| Yes | 4 (66,7%) | 33 (44%) |  |
| **Indisposition, n (%)** |  |  | >0,999 |
| No | 1 (14,3%) | 15 (20%) |  |
| Yes | 6 (85,7%) | 60 (80%) |  |
| **Muscle pain, n (%)** |  |  | >0,999 |
| No | 2 (28,6%) | 22 (29,7%) |  |
| Yes | 5 (71,4%) | 52 (70,3%) |  |
| **Headache, n (%)** |  |  | 0,633 |
| No | 2 (28,6%) | 15 (20,3%) |  |
| Yes | 5 (71,4%) | 59 (79,7%) |  |
| **Sore throat, n (%)** |  |  | >0,999 |
| No | 4 (57,1%) | 42 (56,8%) |  |
| Yes | 3 (42,9%) | 32 (43,2%) |  |
| **Reduction of smell, n (%)** |  |  | 0,433 |
| No | 5 (71,4%) | 35 (50%) |  |
| Yes | 2 (28,6%) | 35 (50%) |  |

Chi-square test; <sup>a</sup> Fisher's exact test; # Likelihood ratio test; <sup>b</sup> t-Student test; £ Mann-Whitney test; SD: standard deviation; min.: minimum; max.: maximum; N.: number.

We can see that patients who followed the protocol treatment for COVID-19 with hydroxychloroquine and azithromycin for 5 days presented improvement of clinical symptoms statistically higher (clinical percentage improvement score) than patients who opted for not following the protocol (p > 0,05).

On Table 7 we compare patients with mild symptoms with patients who didn’t take antibiotics. Those who followed the protocol specific for COVID-19 presented a higher score of clinical percentage improvement after 5 days, with statistical significance (p <0,001). Twelve of the 20 patients with mild symptoms had already initiated antibiotic therapy (with cefalexin, azithromycin or other) before the initial telemedicine consultation and therefore were excluded from this analysis.

**Table 7.**
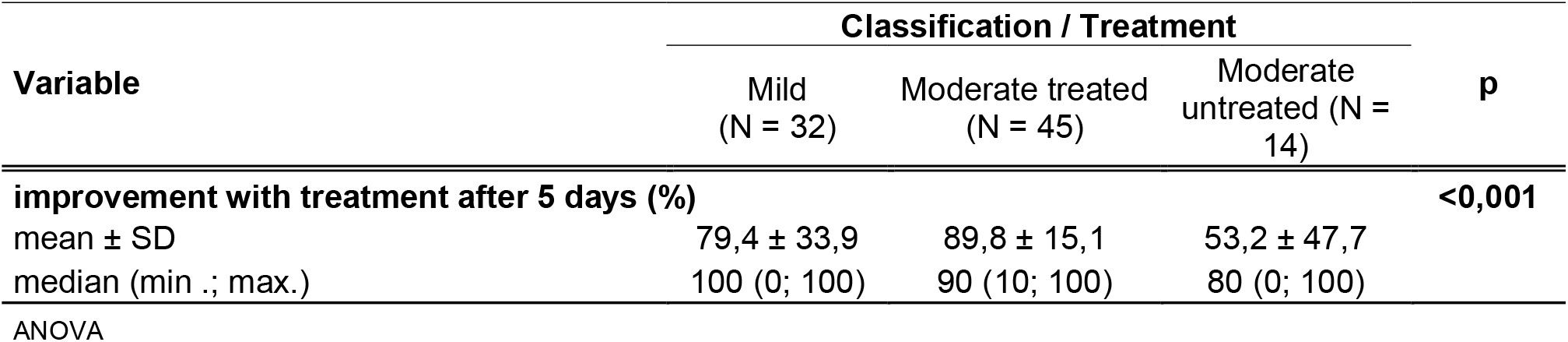
Description of the symptoms improvement, of the patients who followed treatment protocol, after 5 days, and of those with mild symptoms who did not take antibiotic therapy and the result of the comparison. MMC, 2020.

| Variable | Classification / Treatment |  |  | p |
| --- | --- | --- | --- | --- |
|  | Mild<br>(N = 32) | Moderate treated<br>(N = 45) | Moderate<br>untreated (N =<br>14) |  |
| **improvement with treatment after 5 days (%)** |  |  |  | **<0,001** |
| mean ± SD | 79,4 ± 33,9 | 89,8 ± 15,1 | 53,2 ± 47,7 | |
| median (min .; max.) | 100 (0; 100) | 90 (10; 100) | 80 (0; 100) |  |

ANOVA

## Discussion

There are already, in the scientific literature, research that describe the potential use of telemedicine in situations of disasters and public emergencies [1]. Disasters and pandemics represent unique challenges for health services and need fast and efficient attitudes and results to avoid contagion and transmissibility among health professionals. Health services including health maintenance organizations invested in telemedicine and positioning to ensure that COVID-19 patients receive care and orientation without the need of leaving their homes, prescribing medication and guiding to seek hospitals only when truly needed [3,8].

Our results analyzed as a whole points, point to real value of telemedicine in the early management of COVID-19.

On Figure 1 we outline the flow followed in our project. Our study was performed during the first semester of 2020 when the initial therapy of COVID-19 was still predominantly centered on the use of antimalarials, antibiotics, analgesics in an outpatient set up, on the minor number of patients requiring hospitalization O2 therapy, steroids, anticoagulants and mechanical ventilation were up to the doctor team in the respective hospital.

The use of tele screening in all symptomatic patients using the scale mentioned above on the 5^th^ day appeared to be very helpful to the large number of patients, avoiding hospitalization, maintaining virtual face to face and capable of selecting that would need to be referred to hospital for continuous therapy. We have realized the greatest demand for our telemedicine service came from patients already in-home isolation. These patients referred apprehension of getting contaminated and presented several doubts about the necessary care to be taken for their and their families biosafety in this pandemic.

Until mid-April, Brazil’s Health Ministry (HM), the States and Cities made only a few COVID-19 tests available for non-hospitalized population by RT-PCR and quick-test with antibodies (IgG/IgM) measurements. Even for patients in the private services and with good financial conditions, the tests were available only for a few of the hospitalized patients or non-hospitalized patients who would pay the equivalent of almost 1/3 of Brazil’s minimum wage.

After confirmation that the virus was in communitarian sustained transmission, the Health Ministry suggested the diagnosis was made by clinical and epidemiological criteria (identifying the confirmed source case); these were the criteria we used in our research for screening the probable new cases. Of the studied population, 41,8% had been in contact with people who presented flu-like symptoms, 21,4% had been in contact with people who presented positive results por RT-PCR for COVID-19, 91% did not refer recent travel, 9% had recently traveled. The 3 patients in group 4 (severe symptoms), who were immediately referred to a hospital, took the RT-PCR for COVID-19 test and presented positive results. Treatment was evaluated in all symptomatic patients by a clinical improvement scale on the 5^th^ day of therapy.

The possible association between staggered groups was performed, indisposition was the most frequent symptom (p> 0.999), and loss of appetite was increasing with the severity of symptoms (p = 0.670).

April, Brazil’s Health Ministry (HM), the States and Cities made only a few COVID-19 tests available for non-hospitalized population by RT-PCR and quick-test with antibodies (IgG/IgM) measurements. Even for patients in the private services and with good financial conditions, the tests were available only for a few of the hospitalized patients or non-hospitalized patients who would pay the equivalent of almost 1/3 of Brazil’s minimum wage.

On the early period of the pandemic in Brazil the Health Ministry suggested the diagnosis was made by clinical and epidemiological criteria (identifying the confirmed source case); these were the criteria we used in our research for screening the probable new cases. Of the studied population, 41,8% had been in contact with people who presented flu-like symptoms, 21,4% had been in contact with people who presented positive results por RT-PCR for COVID-19, 91% did not refer recent travel, 9% had recently traveled. The 3 patients in group 4 (severe symptoms), who were immediately referred to a hospital, took the RT-PCR for COVID-19 test and presented positive results.

We used an outpatient treatment protocol with hydroxychloroquine and azithromycin after review of the literature made available since the beginning of the pandemic [5,8]. The goal was to evaluate if the patients with moderate symptoms and probable poor prognosis would present less morbimortality in the use of this protocol. The choice of medication was also based on the availability and cost of the medications in Brazilian drug stores and in its wide use in clinical practice for other diseases with safety regarding its collateral effects [6]. During the period we carried out the study the protocol therapy used was the most frequent being used in various centers around the world although controversial evidence of benefit appear to increase during final trimester of 2020.

Our study confirms that telemedicine service is an important strategy for population in pandemic situation, it may be used massively for several health systems around the world, particularly in a country like Brazil with its continental geographic area avoiding the unnecessary direct contact among suspected patients and health care givers, providing quality information and performing screening of the cases. Although we used a standard protocol drug therapy, that was more widespread at the time of the study was performed medications therapy should be adjusted by the medical team performing the practice of telemedicine.

## Data Availability

All data produced in the present study are available upon reasonable request to the authors
All data produced in the present work are contained in the manuscript

## Notes

### Competing Interest Statement

The authors have declared no competing interest.

### Clinical Trial

Clinical trial ID: 30246520.0.0000.0069 and Approval Number: 3.947.259

### Funding Statement

This study did not receive any funding

### Author Declarations

Ethics committee/IRB of CENTRO DE REFERENCIA DA SAUDE DA MULHER - CRSM gave ethical approval for this work. Approval number: 3.947.259

## References

1. Lurie N, Carr BG. The role of telehealth in the medical response to disasters. JAMA Intern Med. 2018;178(6):745–6.

2. AAAAI. https://www.aaaai.org/practice-resources/running-your-practice/practice-management-resources/telemedicine (Accessed on April 21, 2020)

3. Portnoy, J., Waller, M., & Elliott, T. (2020). Telemedicine in the Era of COVID-19. J Allergy Clin Immunol Pract. 2020 May;8(5):1489–1491. doi: 10.1016/j.jaip.2020.03.008. Epub 2020 Mar 24.

4. Hollander, J. E., & Carr, B. G. (2020). Virtually perfect? Telemedicine for covid-19. N Engl J Med 2020; 382:1679–1681

5. Joseph T. Pulmonologist’s Consensus on Covid-19. Int Pulmonologists Consens Covid 19 [Internet]. 2020;2(April):1–72. Available from: https://www.researchgate.net/profile/Tinku_Joseph/publication/340862051_COVID-19_E-Book_International_Pulmonologist’s_consensus_on_COVID-19_-_2nd_Edition/links/5ea150cf299bf143894015e9/COVID-19-E-Book-International-Pulmonologists-consensus-on-COVID-19-2nd

6. Simpson TF, Kovacs RJ, Steckler EC. Ventricular Arrhythmia Risk Due to Hydroxychloroquine-Azithromycin Treatment For COVID-19. Cardiol Mag [Internet]. 2020;(Mar 29):1–9. Available from: https://www.acc.org/latest-in-cardiology/articles/2020/03/27/14/00/ventricular-arrhythmia-risk-due-to-hydroxychloroquine-azithromycin-treatment-for-covid-19

7. Kirkwood, B.R., Sterne, J.A.C., 2006. Essential Medical Statistics, second ed. Blackwell Science, Malden. chapter 35

8. Million M, Lagier JC, Gautret P, Colson P, Fournier PE, Amrane S, et al. Early treatment of COVID-19 patients with hydroxychloroquine and azithromycin: A retrospective analysis of 1061 cases in Marseille, France. Travel Med Infect Dis. 2020 May-Jun;35:101738. doi: 10.1016/j.tmaid.2020.101738. Epub 2020 May 5. PMID: 32387409; PMCID: 6PMC7199729.

