## Supplemental material for "Evaluation of patients treated by telemedicine in the COVID-19 pandemic by a private clinic in São Paulo, Brazil: A non-randomized clinical trial preliminary study"

**SUPPLEMENTAL I**

**Initial Screening Questionnaire**

Name:

WhatsApp number:

Birth Date:

Zip Code:

Address:

Race/ Color:

Marital Status:

Religion:

Number of people living in the same residency:

Number of people living in the same residency with symptoms:

Motive for seeking telemedicine medical care:

Someone else in the family presenting symptoms?

If yes, keep filling the questionnaire:

Fill the name, gender, age, and symptoms of people with symptoms:

Previous medical conditions:

Previous surgeries:

Medications in use:

Smoker?

Alcohol usage?

Drug usage:

Are you feeling well at the moment? Yes or No

Fill yes or no for the following symptoms, when they started and how long they lasted:

1. Date of onset of symptoms:

2. Initial symptom:

3. Fever (quantify temperature):

4. Cough:

5. Nasal congestion:

6. Diarrhea (specify if any bleeding in stool was seen and the number of episodes in a day):

7. Conjunctival irritation:

8. Vomit:

9. Loss of appetite

10. Weight loss (quantify previous and current weight)

11. Did symptoms improve with symptomatic medication (dipyrone or acetaminophen)?

12. Shortness of breath:

13. Indisposition:

14.Muscle pain:

15.Headache:

16. Sore throat:

17. Did you have contact with anyone who was sick?

18. Recent travels? If yes, what was the destination?

19. I declare to agree to participate in the scientific research/clinical trial: yes or no

20. Did you take any laboratory test for COVID-19? If yes, inform the date and result

21. Did you take any treatment directed to COVID-19 (hydroxychloroquine / azithromycin)? Describe in detail the date you started the treatment with the medication and the evolution of your symptoms and if there was any adverse effect:

22.  Did you notice symptoms improvement after bath?

23. Did you present any loss of smell (anosmia)?

**SUPPLEMENTAL II**

**Clinical criteria for definition of mild cases**

- Self-limited infection of upper respiratory tract
- Low fever, sporadic dry cough, rhinorrhea, odynophagia
- Absence of dyspnea
- Conjunctival irritation
- Gastrointestinal symptoms: self-limited nausea / vomit or diarrhea
- Preserved conscious level
- Immunocompetent patients
- Clinical improvement with symptomatic medication

**Clinical criteria for definition of moderate cases**

- Persistent infection of upper respiratory tract
- Recurrent low fever, persistent cough, mild dyspnea, mild muscle pain
- Associated gastrointestinal symptoms: recurrent nausea / vomit or diarrhea
- Preserved conscious level
- Immunocompetent patients with low cardiologic risk for ventricular arrhythmia according to the *American College of Cardiology* (ATTATCHMENT V) score

**Clinical criteria for definition of severe cases**

- Moderate or severe dyspnea
- High persistent fever (>38,5^0^), moderate or intense muscle pain, chill, sweating, important loss of weight
- Increase in respiratory rate > 20 per minute in adults
- Intense and persistente gastrointestinal symptoms
- Compromised conscious level
- Immunodepressed patients or those with moderate or high cardiovascular risk for ventricular arrhythmia according to the *American College of Cardiology* (ATTATCHMENT V) score

**SUPPLEMENTAL III**

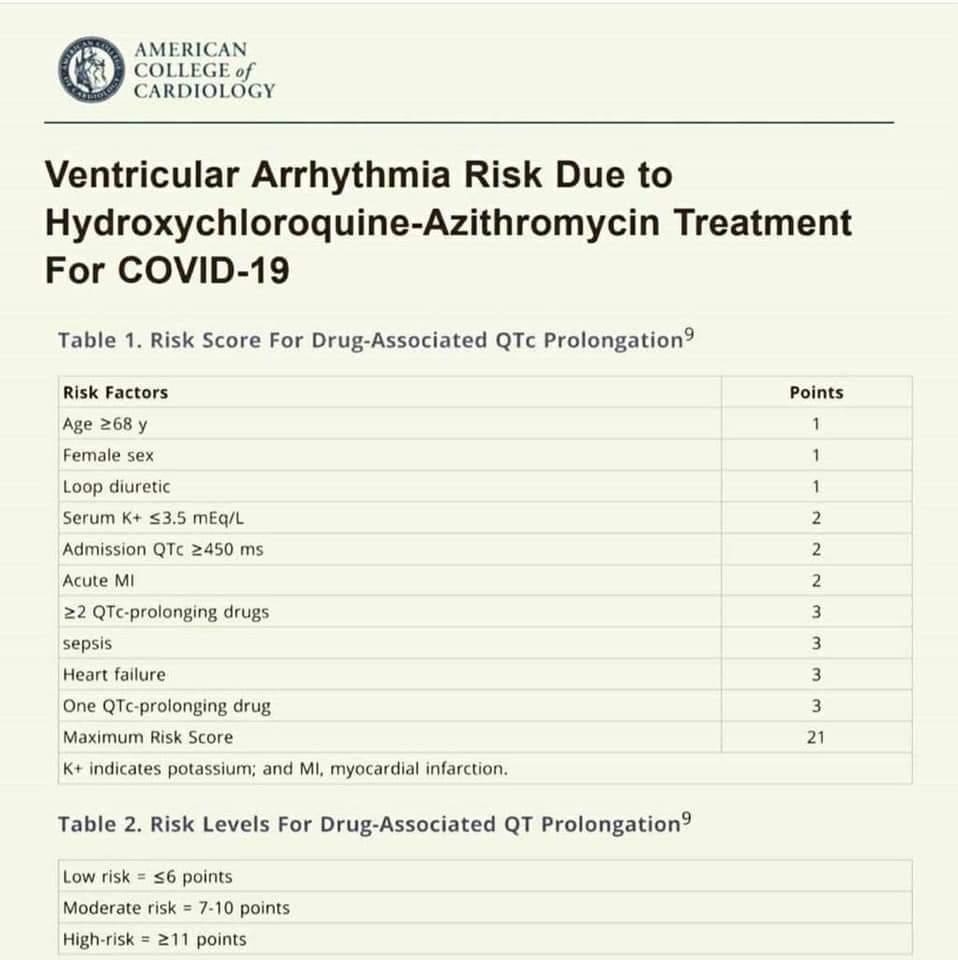
